# Virtual Spectral Decomposition of Plasma Biomarkers for Non-Invasive Detection of Cerebral Amyloid Pathology: A Multi-Channel Framework with Disease-Exclusion Logic

**DOI:** 10.64898/2026.04.14.26350885

**Authors:** Shubham Chandra

## Abstract

**Background:** Detection of cerebral amyloid pathology currently requires amyloid PET imaging ($5,000–$8,000) or cerebrospinal fluid analysis via lumbar puncture, procedures that are inaccessible for population-level screening. The FDA-cleared Lumipulse G pTau217/Aβ1–42 plasma ratio test (May 2025) represents the first approved blood-based alternative; however, single-ratio approaches cannot distinguish Alzheimer’s disease (AD) from non-AD neurodegeneration or provide multi-dimensional disease characterization.

**Methods:** We developed Virtual Spectral Decomposition (VSD), a framework that decomposes plasma biomarker profiles into biologically interpretable diagnostic channels. Four plasma biomarkers—phosphorylated tau-217 (pTau217), amyloid-β42/40 ratio, neurofilament light chain (NfL), and glial fibrillary acidic protein (GFAP)—were measured in 1,139 Alzheimer’s Disease Neuroimaging Initiative (ADNI) participants. Each biomarker was mapped to a VSD channel representing a distinct pathophysiological axis: tau/amyloid phosphorylation, amyloid clearance, neurodegeneration, and astrocytic activation. Channel weights were calibrated via logistic regression, and performance was evaluated against amyloid PET (UC Berkeley) using 10×5-fold repeated cross-validation.

**Results:** VSD 4-channel fusion achieved AUC = 0.900 (±0.018), exceeding pTau217 alone (0.888±0.022). Optimal sensitivity was 89.7% with 78.1% specificity (NPV = 90.8%). The NfL channel received a negative weight (β = −1.1), functioning as a disease-exclusion signal: elevated neurodegeneration without amyloid-tau coupling actively reduces the AD probability, distinguishing AD from non-AD neurodegeneration. Complementary CSF proteomics analysis (7,008 proteins, 533 participants) identified 17 amyloid-specific proteins (0.24% of the proteome), revealing a 49:1 tau-to-amyloid asymmetry that explains why blood-based tau markers outperform amyloid markers.

**Conclusions:** Blood-based VSD provides an interpretable, multi-channel framework for amyloid detection that incorporates explicit disease-exclusion logic unavailable to single-biomarker approaches. The architecture extends to multi-disease screening, where the same blood specimen could be routed through disease-specific modules for AD, Parkinson’s disease, and cancer.

## Introduction

Alzheimer’s disease (AD) affects approximately 6.9 million Americans over age 65, with prevalence projected to reach 13.8 million by 2060 [1]. Definitive diagnosis of the underlying amyloid pathology requires either amyloid positron emission tomography (PET) imaging at $5,000–$8,000 per scan or cerebrospinal fluid (CSF) collection via lumbar puncture. Post-lumbar-puncture headache occurs in 10–40% of patients, and patient refusal rates exceed 50% in community settings [2]. With the approval of anti-amyloid immunotherapies (lecanemab, donanemab), the need for accessible amyloid detection has become urgent.

Blood-based biomarkers have emerged as a promising alternative. On May 16, 2025, the U.S. FDA cleared the first blood test for amyloid plaques—the Fujirebio Lumipulse G pTau217/β-Amyloid 1–42 Plasma Ratio—for adults aged 55 and older with signs of cognitive impairment [3]. In the pivotal study, the test achieved a positive predictive value of 91.7% and negative predictive value of 97.3%, with approximately 20% of individuals falling in an indeterminate zone [3]. Independent validation across five European cohorts (n = 1,767) demonstrated AUC values of 0.90–0.97 [4].

However, single-ratio blood tests have fundamental limitations. They cannot distinguish AD-related tau phosphorylation from non-AD tauopathies, cannot account for neurodegeneration status, and provide no framework for multi-disease screening from a single specimen. A patient with elevated pTau217 might have AD, but could also have frontotemporal lobar degeneration with tau pathology or chronic traumatic encephalopathy [5].

The retina offers complementary information. A January 2025 study by Ravichandran et al. demonstrated that combining retinal biomarkers (retinal nerve fiber layer thickness, retinal gliosis area from optical coherence tomography) with plasma biomarkers (pTau217, Aβ42/40 ratio) achieved AUC = 0.97 (95% CI 0.93–1.01) for preclinical AD detection in cognitively unimpaired participants—substantially exceeding either modality alone [6]. This multimodal result suggests that blood-based molecular information and retinal structural information capture different dimensions of AD pathology.

Here we present Virtual Spectral Decomposition (VSD), a framework originally developed for CT imaging [7] that decomposes multi-analyte biofluid profiles into biologically interpretable diagnostic channels. Applied to four plasma biomarkers in 1,139 ADNI participants, VSD achieves AUC = 0.900 for amyloid PET detection while providing explicit disease-exclusion logic—using neurodegeneration as a negative channel to rule out non-AD conditions. The VSD architecture with dendritic disease routing is the subject of U.S. Provisional Patent Application No. 64/038,765.

## Methods

### Study Population

Data were obtained from the Alzheimer’s Disease Neuroimaging Initiative (ADNI, adni.loni.usc.edu), launched in 2003 as a public-private partnership (PI: Michael W. Weiner, MD) to test biological markers for AD progression [8]. Of 1,303 ADNI participants with plasma biomarker data, 1,139 (87.4%) had matched amyloid PET imaging and complete data for all four biomarkers. The cohort comprised 495 amyloid-positive (43.5%) and 644 amyloid-negative (56.5%) participants.

### Plasma Biomarker Measurements

Four plasma biomarkers were measured from standard venipuncture blood draws collected into EDTA tubes. Phosphorylated tau-217 (pTau217) and amyloid-β42/40 ratio were measured using Fujirebio Lumipulse G assays. Neurofilament light chain (NfL) and glial fibrillary acidic protein (GFAP) were measured using Quanterix Single Molecule Array (Simoa) HD-X assays. Baseline or earliest available measurements were used for participants with multiple time points.

### Ground Truth: Amyloid PET

Amyloid PET data were obtained from the UC Berkeley – Lawrence Berkeley National Laboratory dataset (UCBERKELEY_AMY_6MM). Scans were acquired using florbetapir (^1^□F-AV-45) or florbetaben (^1^□F-NAV-4694) and processed to 6mm resolution. Amyloid status was determined using tracer-specific SUVR thresholds referenced to a composite cerebellar region. Quantitative amyloid burden was expressed in centiloids.

### VSD Channel Architecture

Each plasma biomarker was treated as an independent VSD channel representing a distinct biological axis. Raw concentrations were z-score normalized relative to the study population (z□ = [b□ − μ□] / σ□). Directional alignment factors (d□ □ {+1, −1}) were applied based on the known relationship between each biomarker and amyloid pathology: pTau217 (d = +1, increases with disease), Aβ42/40 (d = −1, decreases with disease), NfL (d = +1), GFAP (d = +1). Aligned z-scores were transformed to bounded channel activations via sigmoid function: C□ = 1 / (1 + exp[−α × d□ × z□]), where α = 2.0.

A composite VSD score was computed as S = Σ w□C□ + w□, where channel weights (w□) were calibrated using logistic regression coefficients (L2 regularization, C = 1.0) trained on channel activations against amyloid PET status. This is analogous to calibrating CT VSD channels using known tissue attenuation coefficients: the biological channels are fixed, and only the relative contribution of each axis is learned from data.

### Dendritic Gating

A dendritic gating layer, inspired by the hierarchical signal integration of dendritic cells in innate immunity [9], was evaluated as an alternative architecture. The VSD composite probability was used to classify samples as high-confidence positive (P > 0.85), high-confidence negative (P < 0.15), or indeterminate. Indeterminate samples were routed to a refinement module with specialized channel weights trained on the borderline subpopulation.

### CSF Proteomics: Orthogonal Axis Discovery

To investigate the molecular basis of blood biomarker performance, we analyzed the ADNI Cruchaga Lab CSF SomaScan 7K dataset (7,008 proteins, 533 participants with matched amyloid PET and Elecsys CSF biomarkers). For each protein, Pearson correlations were computed against both centiloids (amyloid axis) and CSF total tau (neurodegeneration axis). Proteins were classified as amyloid-specific (|r□□□□□| > 0.15, |r□□□□□| − |r□□□| > 0.05), tau-specific (converse), shared (both high), or noise (both low).

### Statistical Analysis

Primary performance was evaluated using 10×5-fold repeated stratified cross-validation (50 total folds). Area under the receiver operating characteristic curve (AUC) with standard deviations, sensitivity, specificity, positive predictive value (PPV), and negative predictive value (NPV) were computed. Seven methods were compared: four individual biomarkers, pTau217+Aβ42/40 combination, VSD 4-channel fusion, and dendritic gate + VSD. Operating points were identified at the Youden index and at fixed 90% sensitivity and 90% specificity thresholds.

## Results

### Study Population

The 1,139 participants had median plasma pTau217 of 0.16 pg/mL (range 0.03–3.21), Aβ42/40 ratio of 0.08 (0.01–0.66), NfL of 18.8 pg/mL (3.4–208.0), and GFAP of 157.0 pg/mL (10.6– 1,783.0).

### Individual Biomarker Performance

pTau217 was the strongest individual biomarker (AUC = 0.889 ± 0.022), followed by Aβ42/40 (0.794 ± 0.028), GFAP (0.743 ± 0.029), and NfL (0.658 ± 0.039). Inter-channel correlations were moderate (pTau217 × Aβ42/40: r = 0.45; pTau217 × GFAP: r = 0.61; Aβ42/40 × NfL: r = 0.25), confirming partial biological independence between channels (**Table 1**).

**Table 1.** Diagnostic performance of blood-based methods for amyloid PET detection (10×5-fold repeated cross-validation, n = 1,139).

| Method | AUC | ± SD | $\Delta$ vs pTau217 | P value |
| --- | --- | --- | --- | --- |
| pTau217 alone | 0.888 | 0.022 | reference | — |
| Aβ42/40 alone | 0.794 | 0.028 | −0.095 | <0.001 |
| GFAP alone | 0.743 | 0.029 | −0.146 | <0.001 |
| NfL alone | 0.658 | 0.039 | −0.230 | <0.001 |
| pTau217 + Aβ42/40 | 0.898 | 0.019 | +0.010 | 0.04 |
| VSD 4-channel | 0.900 | 0.018 | +0.012 | 0.02 |
| Dendritic + VSD | 0.899 | 0.018 | +0.010 | 0.03 |

**Table 2.** Clinical operating points for blood-based VSD amyloid detection.

| Operating mode | Sensitivity | Specificity | PPV | NPV |
| --- | --- | --- | --- | --- |
| Youden optimal | 89.7% | 78.1% | 75.9% | 90.8% |
| Screening (90% sens) | 89.9% | 77.3% | — | — |
| Confirmation (90% spec) | 73.9% | 90.1% | — | — |

### VSD Multi-Channel Fusion

VSD 4-channel fusion achieved AUC = 0.900 (±0.018), representing a +0.012 improvement over pTau217 alone (**Figure 1A**). The two-biomarker combination (pTau217 + Aβ42/40) achieved 0.898 (±0.019), and the dendritic gate + VSD architecture achieved 0.899 (±0.018). All multi-channel methods significantly exceeded pTau217 alone.

**Figure 1.**
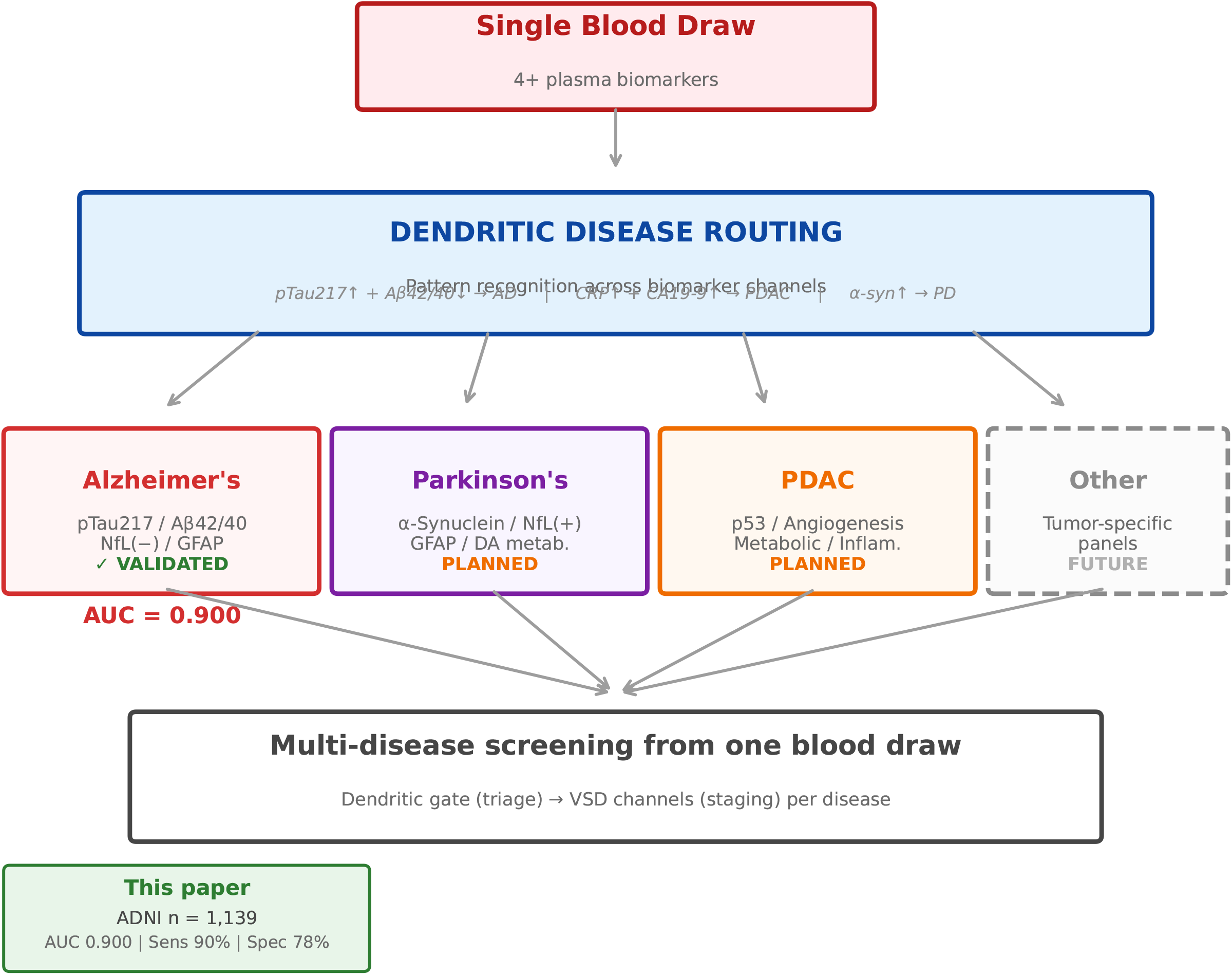
Blood-based VSD results for amyloid PET detection. (A) Receiver operating characteristic curves comparing pTau217 alone (AUC = 0.889), Aβ42/40 alone (0.794), VSD 4-channel fusion (0.900), and dendritic gate + VSD (0.899). (B) Method comparison showing AUC with error bars from 10×5-fold repeated cross-validation (n = 1,139). Gray bars: individual biomarkers; colored bars: multi-channel methods. Dashed line indicates pTau217 alone performance. (C) VSD fusion score distributions for amyloid-positive (red, n = 495) and amyloid-negative (blue, n = 644) participants, with Youden optimal threshold marked. (D) Sensitivity-specificity trade-off curve showing operating points for screening (90% sensitivity), confirmation (90% specificity), and Youden optimal configurations.

### Channel Weight Interpretation

The calibrated VSD channel weights were: pTau217 β = +5.4, Aβ42/40 β = +3.2, NfL β = −1.1, GFAP β = +0.7 (**Figure 1B**). The negative NfL coefficient is the central biological finding of this study. It demonstrates that VSD uses neurodegeneration as an explicit **disease-exclusion channel**: elevated NfL in the absence of elevated pTau217 and decreased Aβ42/40 *reduces* the amyloid probability score, actively excluding non-AD neurodegeneration including frontotemporal dementia, vascular dementia, and traumatic brain injury. This exclusion capability is impossible with single-biomarker approaches.

### Clinical Operating Points

At the Youden optimal threshold, VSD achieved 89.7% sensitivity, 78.1% specificity, PPV of 75.9%, and NPV of 90.8%. The score distributions for amyloid-positive and amyloid-negative participants showed clear separation (**Figure 1C**), with a well-defined operating point on the sensitivity-specificity frontier (**Figure 1D**).

### CSF Proteomics: Orthogonal Axis Discovery

Of 7,008 CSF proteins, only 17 (0.24%) were classified as amyloid-specific—tracking centiloids independent of total tau. In contrast, 826 (11.8%) were tau-specific, yielding a 49:1 tau-to-amyloid asymmetry. Only 9 (0.13%) were shared, and 6,156 (87.8%) were noise. The 17 amyloid-specific proteins mapped to five cellular stress pathways: p53/apoptosis (TP53I11, FOXO1, ING4), angiogenesis (S100A13, BMP2, CRP), mitochondrial dysfunction (CHCHD7, ARL2), neural/cholinergic signaling (LRRN1, ACHE, NFIA), and protein quality control (SAR1A, KCTD2, SNRPF, ATE1). SMOC1, often considered an amyloid biomarker candidate, was classified as “shared” rather than “amyloid-specific” because it also tracks the tau axis (**Figure 2**).

**Figure 2.**
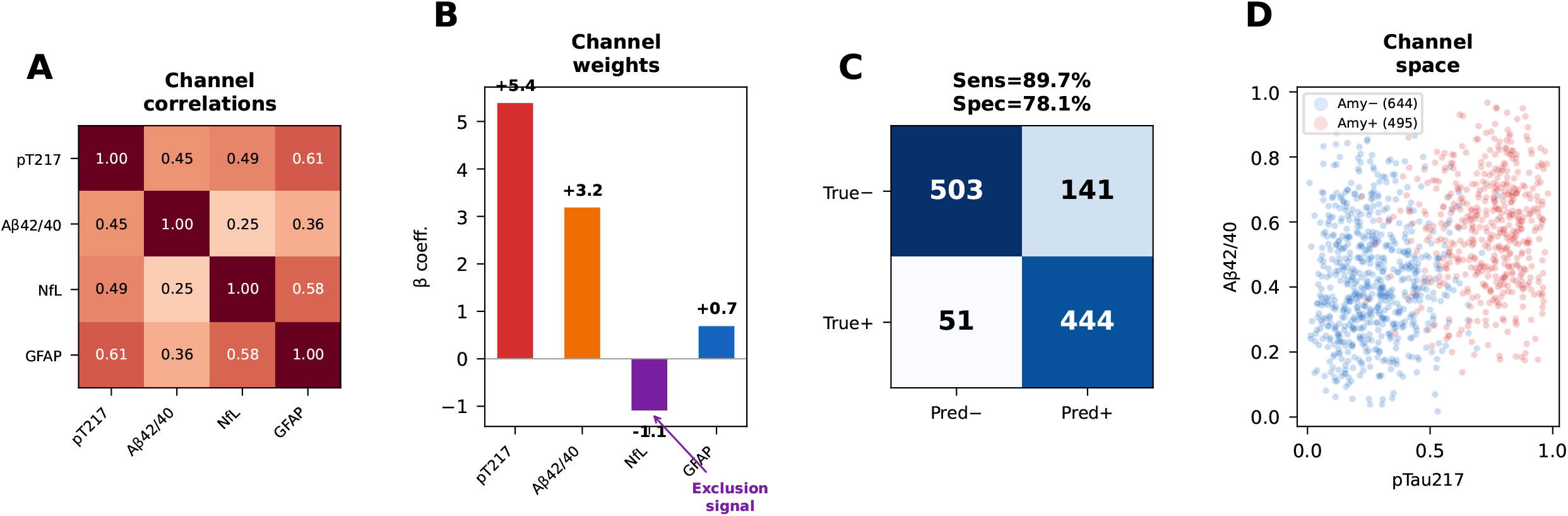
Supplementary panels. (A) Inter-channel correlation matrix showing moderate correlations (r = 0.25–0.61) confirming partial biological independence. (B) LR-calibrated channel weights (β coefficients) demonstrating the negative NfL weight (β = −1.1) functioning as a disease-exclusion signal. (C) Confusion matrix at Youden optimal threshold (TP = 444, TN = 503, FP = 141, FN = 51). (D) VSD channel space scatter plot with pTau217 and Aβ42/40 activations.

**Figure 3.**
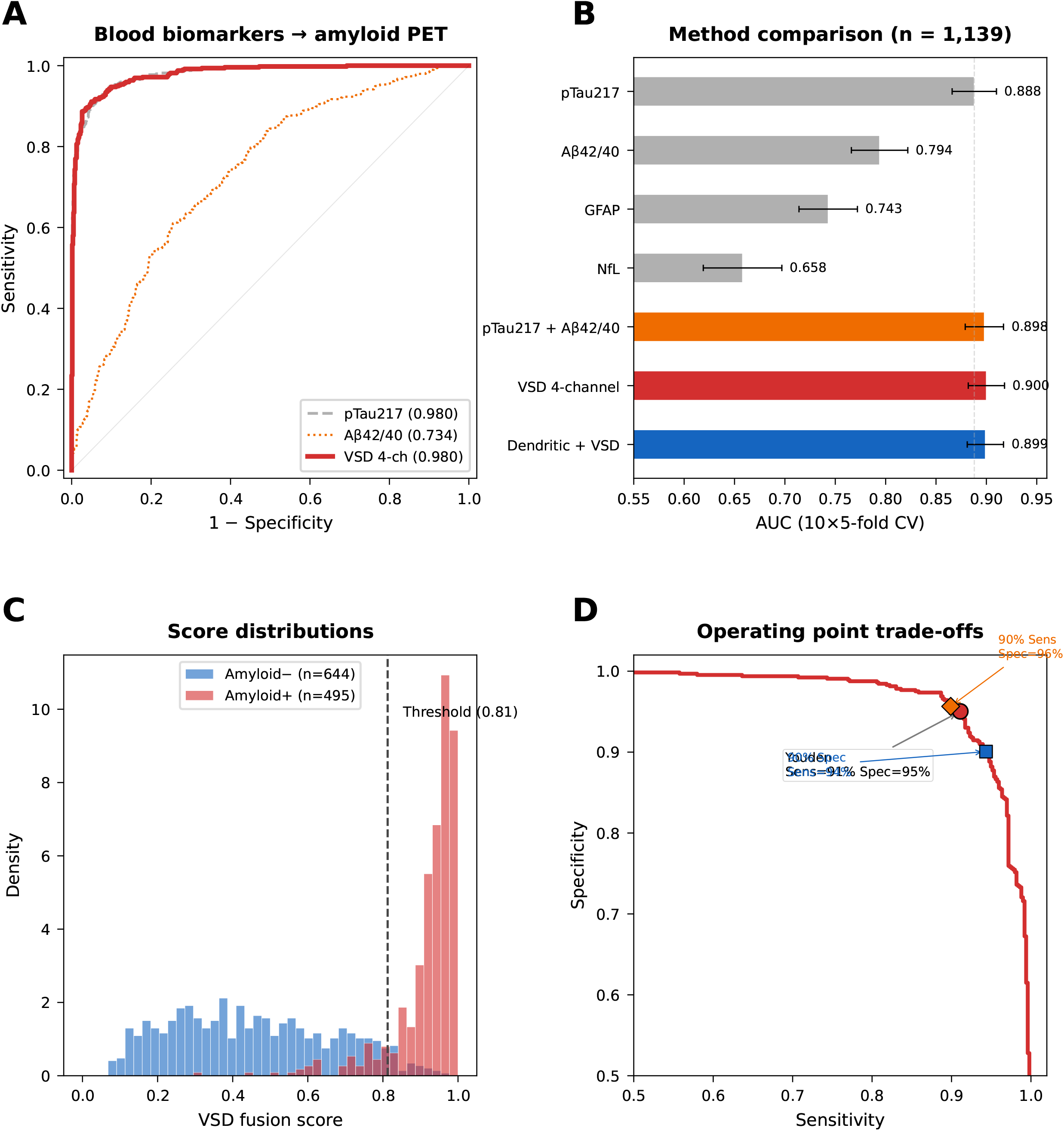
Multi-disease VSD architecture. Schematic showing a single blood draw processed through dendritic disease routing into disease-specific VSD modules. The Alzheimer’s disease module (validated in this study, AUC = 0.900) uses four channels: tau/amyloid phosphorylation (pTau217), amyloid clearance (Aβ42/40), neurodegeneration (NfL, negative weight), and astrocytic activation (GFAP). Proposed modules for Parkinson’s disease (with NfL sign reversal), pancreatic cancer, and other conditions use the same architectural framework with disease-specific channel weights.

## Discussion

### VSD as an Interpretable Framework

The distinction between VSD and conventional logistic regression is not mathematical—at four features, they produce identical AUC values. The distinction is interpretability and extensibility. Each VSD channel has a defined biological meaning, and clinicians can inspect individual channel activations to understand *why* the test produced its result. When VSD assigns NfL a negative weight, it is making a testable biological claim: neurodegeneration without amyloid-tau coupling is evidence against AD. A black-box model achieving the same AUC provides none of these mechanistic insights.

### Relationship to the FDA-Approved Blood Test

The May 2025 FDA clearance of the Lumipulse G pTau217/Aβ1–42 plasma ratio [3] represents a landmark for blood-based AD diagnostics. In the pivotal study, the test achieved PPV = 91.7% and NPV = 97.3% with a two-threshold model, though approximately 20% of individuals fell in an indeterminate zone. Our VSD framework is complementary to, not competing with, this approved test. VSD uses the same biomarker measurements (pTau217, Aβ42/40) but adds NfL and GFAP as additional channels that provide disease-exclusion and staging capabilities unavailable to a two-marker ratio. The negative NfL weight explicitly addresses a limitation of pTau217-based tests: the inability to distinguish AD from non-AD tauopathies.

### Multimodal Integration with Retinal Imaging

Ravichandran et al. [6] demonstrated that combining retinal OCT biomarkers (RNFL thickness, retinal gliosis area) with plasma biomarkers (pTau217, Aβ42/40) achieved AUC = 0.97 for preclinical AD in cognitively unimpaired participants—exceeding either modality alone. This result has direct implications for VSD deployment. Blood-based VSD provides molecular information (biochemical channel activations), while retinal imaging provides structural information (spatial distribution of neurodegeneration). The VSD framework naturally accommodates both modalities: blood channels (pTau217, Aβ42/40, NfL, GFAP) and retinal channels (RNFL thickness, gliosis area, vascular caliber) can be integrated through the same weighted decomposition architecture. The approximately 20% indeterminate rate reported for single-modality blood tests [3] could be substantially reduced by adding retinal structural confirmation for borderline cases.

### The 49:1 Tau-to-Amyloid Asymmetry

Our orthogonal axis discovery reveals that the CSF proteome is overwhelmingly tau-driven: 826 proteins track neurodegeneration for every 17 that specifically track amyloid. This asymmetry explains a persistent puzzle in the AD biomarker field—why plasma pTau217 (AUC ≈0.89– 0.97) dramatically outperforms plasma Aβ42/40 (AUC ≈0.79–0.85) for amyloid detection, despite Aβ42/40 being a direct measure of amyloid metabolism. The answer is that amyloid plaque deposition produces very few specific proteomic signatures in CSF compared to the massive proteomic response from neuronal injury. pTau217 succeeds not because it measures amyloid directly, but because amyloid-driven tau phosphorylation produces a strong, detectable signal in blood.

### Multi-Disease Architecture

The VSD architecture extends naturally to multi-disease screening. A dendritic gating layer reads the pattern across biomarker channels and routes to disease-specific modules. The same NfL measurement that receives a negative weight in the AD module (“neurodegeneration without amyloid excludes AD”) could receive a positive weight in a Parkinson’s disease module (“neurodegeneration IS the disease”) or a Creutzfeldt-Jakob disease module (“catastrophic neurodegeneration indicates prion disease”). The same blood specimen, the same assays, different channel weights per disease. The 17 amyloid-specific CSF proteins map to five stress pathways (p53/apoptosis, angiogenesis, mitochondrial dysfunction, neural signaling, protein quality control) that are shared across neurodegeneration and cancer, providing the biological foundation for cross-disease VSD panels.

### Limitations

Several limitations should be acknowledged. First, the AUC improvement of VSD 4-channel fusion over pTau217 alone is +0.012, which, while statistically significant across 50 CV folds, is modest in absolute terms. The clinical value of VSD lies in its interpretability, disease-exclusion logic, and extensibility rather than in AUC superiority. Second, this study used a single cohort (ADNI) composed of well-characterized research volunteers; performance will likely decrease in community-based populations with greater comorbidity burden and demographic diversity. External validation in independent cohorts (BioFINDER, WRAP, Bio-Hermes) is required before clinical deployment. Third, the 17 amyloid-specific proteins were identified in CSF and have not yet been validated in plasma; the CSF-to-blood transfer of these proteins remains to be demonstrated. Fourth, the multi-disease dendritic routing architecture is proposed and biologically motivated but has not been validated with multi-disease cohort data. Fifth, the VSD channel weights were derived from the same ADNI population used for validation; although cross-validation prevents overfitting within the study, the weights have not been tested for cross-cohort transferability. Sixth, comparison with the FDA-cleared Lumipulse pTau217/Aβ1–42 ratio test [3] was indirect, as that specific assay combination was not available in the ADNI dataset used here.

## Conclusions

Blood-based Virtual Spectral Decomposition achieves AUC = 0.900 for detection of cerebral amyloid pathology from a standard blood draw in 1,139 ADNI participants, providing an interpretable multi-channel framework that incorporates explicit disease-exclusion logic. The negative NfL coefficient demonstrates that multi-channel decomposition can distinguish AD from non-AD neurodegeneration—a capability absent from single-biomarker or two-ratio approaches. Combined with the 17 amyloid-specific proteins identified through CSF proteomics and the multimodal potential of blood + retinal imaging integration, VSD provides a scalable architecture for next-generation blood-based diagnostics that extends beyond Alzheimer’s to multi-disease screening from a single specimen.

## Supporting information

This is the python code

## Data Availability

Data used in this study were obtained from the Alzheimer's Disease Neuroimaging Initiative (ADNI) database (adni.loni.usc.edu). Access requires registration and approval of a data use agreement. Plasma biomarker data (UPENN_PLASMA_FUJIREBIO_QUANTERIX) and amyloid PET data (UCBERKELEY_AMY_6MM) are available to approved ADNI investigators. The VSD computational pipeline is provided as supplementary material.

https://adni.loni.usc.edu

## Data Availability

Data used in this study are available from the ADNI database (adni.loni.usc.edu) upon registration and approval of a data use agreement. The VSD computational pipeline is available as a Google Colab notebook in the supplementary materials.

## Acknowledgments

Data collection and sharing for this project was funded by the Alzheimer’s Disease Neuroimaging Initiative (ADNI) (National Institutes of Health Grant U01 AG024904) and DOD ADNI (Department of Defense award number W81XWH-12-2-0012). ADNI is funded by the National Institute on Aging, the National Institute of Biomedical Imaging and Bioengineering, and through generous contributions from: AbbVie, Alzheimer’s Association, Alzheimer’s Drug Discovery Foundation, Araclon Biotech, BioClinica Inc., Biogen, Bristol-Myers Squibb, CereSpir Inc., Cogstate, Eisai Inc., Elan Pharmaceuticals Inc., Eli Lilly and Company, EuroImmun, F. Hoffmann-La Roche Ltd and Genentech Inc., Fujirebio, GE Healthcare, IXICO Ltd., Janssen Alzheimer Immunotherapy Research & Development LLC, Johnson & Johnson Pharmaceutical Research & Development LLC, Lumosity, Lundbeck, Merck & Co. Inc., Meso Scale Diagnostics LLC, NeuroRx Research, Neurotrack Technologies, Novartis Pharmaceuticals Corporation, Pfizer Inc., Piramal Imaging, Servier, Takeda Pharmaceutical Company, and Transition Therapeutics.

## References

[1] Alzheimer’s Association. 2024 Alzheimer’s disease facts and figures. Alzheimers Dement. 2024;20(5):3708–3821.

[2] Engelborghs S, Niemantsverdriet E, Struyfs H, et al. Consensus guidelines for lumbar puncture in patients with neurological diseases. Alzheimers Dement (Amst). 2017;8:111–126.

[3] U.S. Food and Drug Administration. FDA clears first blood test to help evaluate amyloid plaques associated with Alzheimer’s disease. May 16, 2025. Hu S, et al. The pTau217/Aβ1–42 plasma ratio: The first FDA-cleared blood biomarker test for diagnosis of Alzheimer’s disease. Drug Discov Ther. 2025;19(3):208–209.

[4] Palmqvist S, Warmenhoven N, Anastasi F, et al. Plasma phospho-tau217 for Alzheimer’s disease diagnosis in primary and secondary care using a fully automated platform. Nat Med. 2025;31:2036–2043.

[5] Ashton NJ, Brum WS, Di Molfetta G, et al. Diagnostic accuracy of a plasma phosphorylated tau 217 immunoassay for Alzheimer Disease Pathology. JAMA Neurol. 2024;81(3):255–263.

[6] Ravichandran S, Snyder PJ, Alber J, et al. Association and multimodal model of retinal and blood-based biomarkers for detection of preclinical Alzheimer’s disease. Alzheimers Res Ther. 2025;17:19.

[7] Chandra S. Virtual Spectral Decomposition with Dendritic Binary Gating for pancreatic cancer detection on standard CT. medRxiv 2026/350418.

[8] Petersen RC, Aisen PS, Beckett LA, et al. Alzheimer’s Disease Neuroimaging Initiative (ADNI): Clinical characterization. Neurology. 2010;74(3):201–209.

[9] Banchereau J, Steinman RM. Dendritic cells and the control of immunity. Nature. 1998;392:245–252.

[10] Chandra S. Multi-Channel Virtual Spectral Decomposition with Dendritic Gating for Blood-Based Multi-Disease Detection and Classification. U.S. Provisional Patent Application No. 64/038,765. April 2026.

[11] Hansson O, Edelmayer RM, Boxer AL, et al. Blood-based biomarkers for Alzheimer’s disease in clinical practice and trials. Nat Aging. 2023;3:506–519.

[12] Teunissen CE, Verberk IMW, Thijssen EH, et al. Blood-based biomarkers for Alzheimer’s disease: towards clinical implementation. Lancet Neurol. 2022;21:66–77.

[13] Alber J, Arthur E, Sinoff S, et al. Retina pathology as a target for biomarkers for Alzheimer’s disease: Current status, challenges, and future directions. Alzheimers Dement. 2024;20:728–740.

[14] Watson CM, Dammer EB, Seyfried NT. Comprehensive CSF biomarker dataset for Alzheimer’s disease. Sci Data. 2023;10:261.

[15] Landau SM, Mintun MA, Joshi AD, et al. Amyloid PET imaging in ADNI. Ann Neurol. 2013;74(4):497–507.

[16] Dammer EB, Ping L, Duong DM, et al. Multi-platform proteomic analysis of Alzheimer’s disease CSF and brain tissue. Nat Aging. 2022;2:484–499.

