## Supplementary material for "Virtual Spectral Decomposition of Plasma Biomarkers for Non-Invasive Detection of Cerebral Amyloid Pathology: A Multi-Channel Framework with Disease-Exclusion Logic": This is the python code: VSD_Blood_AD_Complete_Pipeline.py

```
#!/usr/bin/env python3
"""
╔══════════════════════════════════════════════════════════════════════╗
║  BLOOD-BASED VSD FOR ALZHEIMER'S AMYLOID DETECTION                 ║
║  Virtual Spectral Decomposition + Dendritic Gating                 ║
║                                                                     ║
║  Patent Application: 64/038,765                                    ║
║  Inventor: Shubham Chandra | Chandra Associates                    ║
║                                                                     ║
║  Pipeline: Blood Draw → 4 Plasma Proteins → VSD Channels →        ║
║            Dendritic Gate → Amyloid Probability Score              ║
║                                                                     ║
║  Validated: ADNI n=1,139 | AUC=0.900 | Sens=89.7% | Spec=78.1%   ║
║  Ground Truth: UC Berkeley Amyloid PET (Centiloids)                ║
╚══════════════════════════════════════════════════════════════════════╝
"""

# ══════════════════════════════════════════════════════════════════════
# CELL 1: Setup and Configuration
# ══════════════════════════════════════════════════════════════════════
# %% [markdown]
# # Blood-Based VSD for Alzheimer's Amyloid Detection
# **Patent Application No. 64/038,765**
#
# This notebook implements the complete pipeline from raw ADNI data
# to clinical diagnostic output. Each cell corresponds to one step
# in the clinical workflow:
#
# 1. **Setup** — Mount Drive, install dependencies
# 2. **Load Data** — Plasma biomarkers + Amyloid PET ground truth
# 3. **Preprocess** — Quality control, matching, normalization
# 4. **VSD Channels** — Build 4 biological channel activations
# 5. **Calibration** — Learn channel weights via logistic regression
# 6. **Validation** — 10×5-fold cross-validation with full metrics
# 7. **Clinical Report** — Generate diagnostic output for a patient
# 8. **Manuscript Figures** — Publication-quality plots

# %%
# ── Cell 1: Setup ───────────────────────────────────────────────────
from google.colab import drive
drive.mount('/content/drive')

import numpy as np, pandas as pd, warnings, json, os
from sklearn.model_selection import StratifiedKFold, RepeatedStratifiedKFold
from sklearn.metrics import (roc_auc_score, roc_curve, confusion_matrix,
                             classification_report)
from sklearn.linear_model import LogisticRegression
from sklearn.preprocessing import StandardScaler
from scipy.stats import pearsonr
import matplotlib
import matplotlib.pyplot as plt
import matplotlib.patches as mpatches
matplotlib.rcParams.update({
    'figure.dpi': 150, 'font.size': 9, 'font.family': 'sans-serif',
    'axes.linewidth': 0.5, 'savefig.dpi': 300,
})
warnings.filterwarnings('ignore')

# ── Paths (adjust to your Google Drive structure) ───────────────────
BASE_DIR = '/content/drive/MyDrive/VSDbasedBloodBiomarkers'
SOMA_DIR = f'{BASE_DIR}/Somascan'
MERGE_DIR = f'{SOMA_DIR}/ADNIMERGE_extracted/ADNIMERGE2/data'
OUT_DIR = f'{SOMA_DIR}/ADNI_VSD_Results'
os.makedirs(OUT_DIR, exist_ok=True)

# Amyloid PET file — update filename if different
AMY_PET_FILE = f'{SOMA_DIR}/UCBERKELEY_AMY_6MM_14Apr2026.csv'

print('✓ Setup complete')
print(f'  Output directory: {OUT_DIR}')


# ══════════════════════════════════════════════════════════════════════
# CELL 2: Load Raw Data
# ══════════════════════════════════════════════════════════════════════
# %% [markdown]
# ## Step 1: Blood Draw → Measure Proteins
# Load actual plasma protein concentrations measured from ADNI
# participants' blood draws via Fujirebio Lumipulse and Quanterix Simoa.

# %%
import pyreadr

print('═' * 70)
print('STEP 1: LOADING RAW BLOOD DATA')
print('═' * 70)

# ── Load plasma biomarker measurements ──────────────────────────────
# Source: UPenn Fujirebio + Quanterix immunoassays on ADNI plasma
plasma_rda = pyreadr.read_r(f'{MERGE_DIR}/UPENN_PLASMA_FUJIREBIO_QUANTERIX.rda')
plasma_raw = list(plasma_rda.values())[0]

print(f'\nPlasma biomarker dataset: {plasma_raw.shape[0]} measurements')
print(f'  Columns: {list(plasma_raw.columns)}')
print(f'  Assay platforms:')
print(f'    Fujirebio Lumipulse: pTau217, Aβ42, Aβ40')
print(f'    Quanterix Simoa HD-X: NfL, GFAP')

# ── Define the 4 VSD channel biomarkers ─────────────────────────────
CHANNEL_CONFIG = {
    'pT217': {
        'column': 'pT217_F',
        'full_name': 'Phosphorylated Tau-217',
        'assay': 'Fujirebio Lumipulse',
        'units': 'pg/mL',
        'biology': 'Tau phosphorylation driven by amyloid plaques',
        'vsd_channel': 'Tau/Amyloid Phosphorylation',
        'direction': +1,  # increases with disease
        'color': '#D32F2F',
    },
    'AB42_40': {
        'column': 'AB42_AB40_F',
        'full_name': 'Amyloid-β 42/40 Ratio',
        'assay': 'Fujirebio Lumipulse',
        'units': 'ratio',
        'biology': 'Amyloid clearance failure (Aβ42 sequestered into plaques)',
        'vsd_channel': 'Amyloid Clearance',
        'direction': -1,  # DECREASES with disease (inverted)
        'color': '#EF6C00',
    },
    'NfL': {
        'column': 'NfL_Q',
        'full_name': 'Neurofilament Light Chain',
        'assay': 'Quanterix Simoa HD-X',
        'units': 'pg/mL',
        'biology': 'Axonal neurodegeneration (non-specific)',
        'vsd_channel': 'Neurodegeneration',
        'direction': +1,  # increases with damage
        'color': '#7B1FA2',
    },
    'GFAP': {
        'column': 'GFAP_Q',
        'full_name': 'Glial Fibrillary Acidic Protein',
        'assay': 'Quanterix Simoa HD-X',
        'units': 'pg/mL',
        'biology': 'Astrocytic activation and neuroinflammation',
        'vsd_channel': 'Astrocytic Activation',
        'direction': +1,  # increases with activation
        'color': '#1565C0',
    },
}

FEATURE_NAMES = list(CHANNEL_CONFIG.keys())
FEATURE_COLS = [CHANNEL_CONFIG[f]['column'] for f in FEATURE_NAMES]

# ── Get baseline measurement per participant ────────────────────────
plasma_valid = plasma_raw.dropna(subset=['pT217_F', 'NfL_Q'])
if 'VISCODE' in plasma_valid.columns:
    plasma_valid = plasma_valid.copy()
    plasma_valid['is_bl'] = plasma_valid['VISCODE'].astype(str).str.contains(
        'bl|sc', case=False, na=False).astype(int)
    plasma_valid = plasma_valid.sort_values(['RID', 'is_bl'], ascending=[True, False])
plasma_bl = plasma_valid.drop_duplicates('RID', keep='first')

print(f'\n  Unique participants with plasma data: {len(plasma_bl)}')
for name, cfg in CHANNEL_CONFIG.items():
    vals = plasma_bl[cfg['column']].dropna()
    print(f'  {name:8s} ({cfg["column"]:12s}): n={len(vals)}, '
          f'median={vals.median():.2f}, range=[{vals.min():.2f}, {vals.max():.2f}]')

# ── Load amyloid PET ground truth ───────────────────────────────────
print(f'\n  Loading amyloid PET: {AMY_PET_FILE}')
amy_pet = pd.read_csv(AMY_PET_FILE)
print(f'  Amyloid PET scans: {len(amy_pet)}')

# Baseline PET per participant
if 'VISCODE2' in amy_pet.columns:
    amy_pet_s = amy_pet.copy()
    amy_pet_s['is_bl'] = amy_pet_s['VISCODE2'].astype(str).str.contains(
        'bl|sc', case=False, na=False).astype(int)
    amy_pet_s = amy_pet_s.sort_values(['RID', 'is_bl'], ascending=[True, False])
else:
    amy_pet_s = amy_pet.copy()
amy_bl = amy_pet_s.drop_duplicates('RID', keep='first')
print(f'  Unique participants with PET: {len(amy_bl)}')

print('\n✓ Raw data loaded')


# ══════════════════════════════════════════════════════════════════════
# CELL 3: Preprocess — Match Plasma to PET, Quality Control
# ══════════════════════════════════════════════════════════════════════
# %% [markdown]
# ## Step 2: Spin the Blood → Extract Plasma → Match to Ground Truth
# Centrifuge separates plasma from cells. We match plasma protein
# measurements to amyloid PET results for the same participants.

# %%
print('═' * 70)
print('STEP 2: PREPROCESSING AND MATCHING')
print('═' * 70)

# ── Match plasma to PET by participant RID ──────────────────────────
plasma_rids = set(plasma_bl['RID'].astype(int).values)
pet_rids = set(amy_bl['RID'].astype(int).values)
matched_rids = sorted(plasma_rids & pet_rids)
print(f'\n  Plasma participants: {len(plasma_rids)}')
print(f'  PET participants: {len(pet_rids)}')
print(f'  Matched (both): {len(matched_rids)}')

# ── Build matched dataset ───────────────────────────────────────────
rows = []
for rid in matched_rids:
    prow = plasma_bl[plasma_bl['RID'].astype(int) == rid].iloc[0]
    arow = amy_bl[amy_bl['RID'].astype(int) == rid].iloc[0]

    entry = {'RID': int(rid)}
    # Plasma biomarkers
    for name in FEATURE_NAMES:
        col = CHANNEL_CONFIG[name]['column']
        entry[name] = float(prow[col]) if pd.notna(prow[col]) else np.nan
    # Amyloid PET
    if 'AMYLOID_STATUS' in arow.index and pd.notna(arow['AMYLOID_STATUS']):
        entry['amy_status'] = int(arow['AMYLOID_STATUS'])
    else:
        entry['amy_status'] = np.nan
    if 'CENTILOIDS' in arow.index and pd.notna(arow['CENTILOIDS']):
        entry['centiloids'] = float(arow['CENTILOIDS'])
    rows.append(entry)

df = pd.DataFrame(rows)

# ── Filter to complete cases ────────────────────────────────────────
complete = df[FEATURE_NAMES + ['amy_status']].notna().all(axis=1)
df_valid = df[complete].copy().reset_index(drop=True)

n_pos = (df_valid['amy_status'] == 1).sum()
n_neg = (df_valid['amy_status'] == 0).sum()
N = len(df_valid)

print(f'\n  Complete cases: {N}')
print(f'  Amyloid Positive (PET+): {n_pos} ({n_pos/N:.1%})')
print(f'  Amyloid Negative (PET−): {n_neg} ({n_neg/N:.1%})')

# ── Extract arrays ──────────────────────────────────────────────────
X_raw = df_valid[FEATURE_NAMES].values.astype(float)
y = df_valid['amy_status'].values.astype(int)

# ── Z-score normalization ───────────────────────────────────────────
# Store population statistics as calibration constants
scaler = StandardScaler()
X_z = scaler.fit_transform(X_raw)

CALIBRATION = {}
for i, name in enumerate(FEATURE_NAMES):
    CALIBRATION[name] = {
        'mean': float(scaler.mean_[i]),
        'std': float(scaler.scale_[i]),
        'direction': CHANNEL_CONFIG[name]['direction'],
    }
    print(f'  {name:8s}: μ = {scaler.mean_[i]:.4f}, σ = {scaler.scale_[i]:.4f}, '
          f'direction = {"↑" if CHANNEL_CONFIG[name]["direction"] == 1 else "↓"}')

print('\n✓ Preprocessing complete')


# ══════════════════════════════════════════════════════════════════════
# CELL 4: Build VSD Channels
# ══════════════════════════════════════════════════════════════════════
# %% [markdown]
# ## Step 3: Feed the Numbers into VSD
# Transform normalized concentrations into bounded channel activations
# via sigmoid function. Each channel represents a biological axis.

# %%
print('═' * 70)
print('STEP 3: VSD CHANNEL COMPUTATION')
print('═' * 70)

# ── VSD Sigmoid Function ────────────────────────────────────────────
# C_i = σ(α × d_i × z_i) = 1 / (1 + exp(-α × d_i × z_i))
# where:
#   z_i = z-score normalized biomarker concentration
#   d_i = direction (+1 or -1)
#   α = steepness parameter (default 2.0)
#   C_i = channel activation ∈ [0, 1]

ALPHA = 2.0  # Sigmoid steepness

def vsd_sigmoid(z, direction, alpha=ALPHA):
    """Transform z-score to bounded channel activation."""
    return 1.0 / (1.0 + np.exp(-alpha * direction * z))

# ── Compute channel activations for all samples ─────────────────────
Ca = np.zeros((N, 4))
for ci, name in enumerate(FEATURE_NAMES):
    d = CHANNEL_CONFIG[name]['direction']
    Ca[:, ci] = vsd_sigmoid(X_z[:, ci], d)

print(f'\n  Sigmoid steepness α = {ALPHA}')
print(f'  Channel activations computed for {N} samples × 4 channels\n')

# ── Per-channel diagnostic performance ──────────────────────────────
print(f'  {"Channel":<25s} {"VSD Axis":<25s} {"AUC":>6s} {"Direction":>10s}')
print(f'  {"─"*70}')
ch_aucs = {}
for ci, name in enumerate(FEATURE_NAMES):
    auc = roc_auc_score(y, Ca[:, ci])
    if auc < 0.5: auc = 1 - auc
    ch_aucs[name] = auc
    cfg = CHANNEL_CONFIG[name]
    print(f'  {cfg["full_name"]:<25s} {cfg["vsd_channel"]:<25s} {auc:>6.3f} '
          f'{"↑ with AD" if cfg["direction"]==1 else "↓ with AD":>10s}')

# ── Channel correlations (orthogonality check) ──────────────────────
print(f'\n  Inter-channel correlations (lower = more orthogonal):')
for i in range(4):
    for j in range(i+1, 4):
        r = pearsonr(Ca[:, i], Ca[:, j])[0]
        orth = '✓ orthogonal' if abs(r) < 0.4 else '~ moderate' if abs(r) < 0.6 else '! correlated'
        print(f'    {FEATURE_NAMES[i]:8s} × {FEATURE_NAMES[j]:8s}: r = {r:+.3f}  {orth}')

print('\n✓ VSD channels computed')


# ══════════════════════════════════════════════════════════════════════
# CELL 5: Calibrate Channel Weights
# ══════════════════════════════════════════════════════════════════════
# %% [markdown]
# ## Step 4: Compute the VSD Score
# Learn optimal channel weights via logistic regression.
# Score = Σ w_i × C_i + w₀
# The channels are fixed biological axes. Only the weights are learned.

# %%
print('═' * 70)
print('STEP 4: CHANNEL WEIGHT CALIBRATION')
print('═' * 70)

# ── Fit logistic regression on full dataset for weight inspection ───
# (Cross-validated weights are computed in Cell 6)
lr_full = LogisticRegression(C=1.0, max_iter=1000, random_state=42)
lr_full.fit(Ca, y)

print(f'\n  VSD Composite Score = ', end='')
terms = []
for ci, name in enumerate(FEATURE_NAMES):
    coef = lr_full.coef_[0][ci]
    sign = '+' if coef > 0 else ''
    terms.append(f'{sign}{coef:.1f} × {name}')
intercept = lr_full.intercept_[0]
sign_i = '+' if intercept > 0 else ''
print(f'{" ".join(terms)} {sign_i}{intercept:.1f}')

print(f'\n  Channel weights (β coefficients):')
for ci, name in enumerate(FEATURE_NAMES):
    coef = lr_full.coef_[0][ci]
    cfg = CHANNEL_CONFIG[name]
    role = 'POSITIVE (disease indicator)' if coef > 0 else 'NEGATIVE (disease EXCLUSION)'
    print(f'    {name:8s}: β = {coef:+.3f}  [{role}]')
    print(f'             {cfg["biology"]}')

print(f'    Intercept: {lr_full.intercept_[0]:+.3f}')

# ── Full-data AUC ───────────────────────────────────────────────────
vsd_scores_full = lr_full.predict_proba(Ca)[:, 1]
auc_full = roc_auc_score(y, vsd_scores_full)
print(f'\n  Full-data AUC: {auc_full:.3f}')

# ── Optimal operating point (Youden) ────────────────────────────────
fpr_f, tpr_f, thr_f = roc_curve(y, vsd_scores_full)
j_idx = np.argmax(tpr_f - fpr_f)
OPT_THRESHOLD = thr_f[j_idx]
OPT_SENS = tpr_f[j_idx]
OPT_SPEC = 1 - fpr_f[j_idx]

preds_opt = (vsd_scores_full >= OPT_THRESHOLD).astype(int)
tn, fp, fn, tp = confusion_matrix(y, preds_opt).ravel()
PPV = tp / (tp + fp)
NPV = tn / (tn + fn)

print(f'\n  Optimal operating point (Youden J):')
print(f'    Threshold: {OPT_THRESHOLD:.4f}')
print(f'    Sensitivity: {OPT_SENS:.1%}')
print(f'    Specificity: {OPT_SPEC:.1%}')
print(f'    PPV: {PPV:.1%}  |  NPV: {NPV:.1%}')
print(f'    TP={tp}  FP={fp}  FN={fn}  TN={tn}')

print('\n✓ Calibration complete')


# ══════════════════════════════════════════════════════════════════════
# CELL 6: Cross-Validation (Rigorous)
# ══════════════════════════════════════════════════════════════════════
# %% [markdown]
# ## Step 5: Validate the Pipeline
# 10×5-fold repeated stratified cross-validation.
# 50 total train/test splits. No data leakage.

# %%
print('═' * 70)
print('STEP 5: 10×5-FOLD CROSS-VALIDATION')
print(f'{N} participants | {n_pos} Amy+ | {n_neg} Amy−')
print('═' * 70)

rskf = RepeatedStratifiedKFold(n_splits=5, n_repeats=10, random_state=42)

cv_results = {name: [] for name in FEATURE_NAMES}
cv_results['VSD_fusion'] = []
cv_results['Dendritic_VSD'] = []
cv_results['pT217_AB42_40'] = []

CONF_THR = 0.15  # Dendritic confidence threshold

for fold_i, (tr, te) in enumerate(rskf.split(Ca, y)):
    Ca_tr, Ca_te = Ca[tr], Ca[te]
    y_tr, y_te = y[tr], y[te]

    # ── Single biomarkers ───────────────────────────────────────────
    for ci, name in enumerate(FEATURE_NAMES):
        a = roc_auc_score(y_te, Ca_te[:, ci])
        cv_results[name].append(a if a > 0.5 else 1 - a)

    # ── pTau217 + Aβ42/40 (two-marker) ─────────────────────────────
    lr2 = LogisticRegression(C=1.0, max_iter=1000, random_state=42)
    lr2.fit(Ca_tr[:, :2], y_tr)
    cv_results['pT217_AB42_40'].append(roc_auc_score(y_te, lr2.predict_proba(Ca_te[:, :2])[:, 1]))

    # ── VSD 4-channel fusion (LR-calibrated) ────────────────────────
    lr4 = LogisticRegression(C=1.0, max_iter=1000, random_state=42)
    lr4.fit(Ca_tr, y_tr)
    vsd_scores = lr4.predict_proba(Ca_te)[:, 1]
    cv_results['VSD_fusion'].append(roc_auc_score(y_te, vsd_scores))

    # ── Dendritic gate + VSD refinement ─────────────────────────────
    probs_tr = lr4.predict_proba(Ca_tr)[:, 1]
    probs_te = vsd_scores.copy()
    final_scores = probs_te.copy()

    # Identify uncertain samples
    uncertain_te = (probs_te > CONF_THR) & (probs_te < (1 - CONF_THR))
    uncertain_tr = (probs_tr > CONF_THR) & (probs_tr < (1 - CONF_THR))

    if uncertain_te.sum() > 5 and uncertain_tr.sum() > 15:
        if len(np.unique(y_tr[uncertain_tr])) == 2:
            lr_ref = LogisticRegression(C=0.5, max_iter=1000, random_state=42)
            lr_ref.fit(Ca_tr[uncertain_tr], y_tr[uncertain_tr])
            final_scores[uncertain_te] = lr_ref.predict_proba(Ca_te[uncertain_te])[:, 1]

    cv_results['Dendritic_VSD'].append(roc_auc_score(y_te, final_scores))

# ── Summary ─────────────────────────────────────────────────────────
method_labels = {
    'pT217': 'pTau217 alone',
    'AB42_40': 'Aβ42/40 alone',
    'NfL': 'NfL alone',
    'GFAP': 'GFAP alone',
    'pT217_AB42_40': 'pTau217 + Aβ42/40',
    'VSD_fusion': 'VSD 4-channel fusion',
    'Dendritic_VSD': 'Dendritic gate + VSD',
}

method_order = ['pT217', 'AB42_40', 'GFAP', 'NfL',
                'pT217_AB42_40', 'VSD_fusion', 'Dendritic_VSD']

print(f'\n  {"Method":<28s} {"AUC":>8s} {"± SD":>8s} {"vs pT217":>10s}')
print(f'  {"─"*56}')
ptau_mean = np.mean(cv_results['pT217'])
summary = {}
for method in method_order:
    vals = cv_results[method]
    m, s = np.mean(vals), np.std(vals)
    delta = m - ptau_mean
    summary[method] = {'mean': m, 'std': s}
    marker = '↑' if delta > 0.002 else '↓' if delta < -0.002 else '='
    print(f'  {method_labels[method]:<28s} {m:>8.3f} {s:>8.3f} {delta:>+9.3f} {marker}')

best = max(summary.items(), key=lambda x: x[1]['mean'])
print(f'\n  ★ BEST: {method_labels[best[0]]} (AUC = {best[1]["mean"]:.3f})')

print('\n✓ Cross-validation complete')


# ══════════════════════════════════════════════════════════════════════
# CELL 7: Clinical Report Generator
# ══════════════════════════════════════════════════════════════════════
# %% [markdown]
# ## Step 6: Generate Clinical Diagnostic Report
# Enter a patient's 4 plasma protein concentrations and get
# a VSD amyloid probability score with clinical interpretation.

# %%
print('═' * 70)
print('STEP 6: CLINICAL REPORT GENERATOR')
print('═' * 70)

def generate_clinical_report(pt217_pgml, ab42_40_ratio, nfl_pgml, gfap_pgml,
                              patient_name="Sample Patient", patient_age=None):
    """
    Generate a VSD clinical diagnostic report from 4 plasma concentrations.

    Parameters:
    -----------
    pt217_pgml : float   - Plasma pTau217 concentration (pg/mL)
    ab42_40_ratio : float - Plasma Aβ42/40 ratio
    nfl_pgml : float     - Plasma NfL concentration (pg/mL)
    gfap_pgml : float    - Plasma GFAP concentration (pg/mL)
    patient_name : str   - Patient identifier
    patient_age : int    - Patient age (optional)
    """

    raw_values = np.array([pt217_pgml, ab42_40_ratio, nfl_pgml, gfap_pgml])

    # Step 1: Z-score normalization using stored calibration constants
    z_scores = np.zeros(4)
    for i, name in enumerate(FEATURE_NAMES):
        cal = CALIBRATION[name]
        z_scores[i] = (raw_values[i] - cal['mean']) / cal['std']

    # Step 2: Directional alignment + sigmoid transformation
    activations = np.zeros(4)
    for i, name in enumerate(FEATURE_NAMES):
        d = CALIBRATION[name]['direction']
        activations[i] = vsd_sigmoid(z_scores[i], d)

    # Step 3: Compute VSD composite score
    probability = lr_full.predict_proba(activations.reshape(1, -1))[0, 1]

    # Step 4: Classification
    if probability > (1 - CONF_THR):
        classification = "AMYLOID POSITIVE"
        confidence = "HIGH"
        color_code = "🔴"
    elif probability < CONF_THR:
        classification = "AMYLOID NEGATIVE"
        confidence = "HIGH"
        color_code = "🟢"
    else:
        classification = "INDETERMINATE"
        confidence = "LOW — recommend retinal imaging or PET confirmation"
        color_code = "🟡"

    # Step 5: Clinical interpretation
    interpretations = []
    if activations[0] > 0.7:
        interpretations.append("Elevated tau phosphorylation consistent with amyloid-driven pathology")
    if activations[1] > 0.6:
        interpretations.append("Impaired amyloid clearance (low Aβ42/40 ratio)")
    if activations[2] > 0.6 and activations[0] < 0.5:
        interpretations.append("⚠ Elevated neurodegeneration WITHOUT amyloid-tau coupling — "
                              "consider non-AD etiology (FTD, vascular, TBI)")
    elif activations[2] > 0.6 and activations[0] > 0.7:
        interpretations.append("Neurodegeneration proportionate to amyloid-tau signal")
    if activations[3] > 0.6:
        interpretations.append("Astrocytic activation detected")

    # ── Print report ────────────────────────────────────────────────
    print(f'\n{"╔" + "═"*66 + "╗"}')
    print(f'{"║"} {"VSD BLOOD-BASED AMYLOID SCREENING REPORT":^64s} {"║"}')
    print(f'{"║"} {"Patent Application No. 64/038,765":^64s} {"║"}')
    print(f'{"╠" + "═"*66 + "╣"}')
    print(f'{"║"} Patient: {patient_name:<55s} {"║"}')
    if patient_age:
        print(f'{"║"} Age: {patient_age:<59d} {"║"}')
    print(f'{"║"} Date: {"April 14, 2026":<55s} {"║"}')
    print(f'{"╠" + "═"*66 + "╣"}')
    print(f'{"║"} {"PLASMA MEASUREMENTS":^64s} {"║"}')
    print(f'{"╟" + "─"*66 + "╢"}')

    for i, name in enumerate(FEATURE_NAMES):
        cfg = CHANNEL_CONFIG[name]
        bar_len = int(activations[i] * 20)
        bar = '█' * bar_len + '░' * (20 - bar_len)
        level = 'HIGH' if activations[i] > 0.7 else 'LOW' if activations[i] < 0.3 else 'MOD'
        print(f'{"║"}  {cfg["full_name"]:<22s} {raw_values[i]:>8.2f} {cfg["units"]:<6s} '
              f'{bar} {level:>4s} {"║"}')

    print(f'{"╠" + "═"*66 + "╣"}')
    print(f'{"║"} {"VSD CHANNEL ACTIVATIONS":^64s} {"║"}')
    print(f'{"╟" + "─"*66 + "╢"}')

    for i, name in enumerate(FEATURE_NAMES):
        cfg = CHANNEL_CONFIG[name]
        print(f'{"║"}  {cfg["vsd_channel"]:<28s} C = {activations[i]:.3f}  '
              f'(β = {lr_full.coef_[0][i]:+.1f})          {"║"}')

    print(f'{"╠" + "═"*66 + "╣"}')
    print(f'{"║"} {"COMPOSITE VSD SCORE":^64s} {"║"}')
    print(f'{"╟" + "─"*66 + "╢"}')
    print(f'{"║"}  {color_code} Amyloid Probability: {probability:.1%:<44s} {"║"}')
    print(f'{"║"}  Classification: {classification:<47s} {"║"}')
    print(f'{"║"}  Confidence: {confidence:<51s} {"║"}')

    print(f'{"╠" + "═"*66 + "╣"}')
    print(f'{"║"} {"CLINICAL INTERPRETATION":^64s} {"║"}')
    print(f'{"╟" + "─"*66 + "╢"}')
    for interp in interpretations:
        # Word wrap
        words = interp.split()
        line = ''
        for word in words:
            if len(line) + len(word) + 1 > 62:
                print(f'{"║"}  {line:<62s}  {"║"}')
                line = word
            else:
                line = f'{line} {word}'.strip()
        if line:
            print(f'{"║"}  {line:<62s}  {"║"}')

    print(f'{"╠" + "═"*66 + "╣"}')
    print(f'{"║"} {"RECOMMENDATION":^64s} {"║"}')
    print(f'{"╟" + "─"*66 + "╢"}')
    if classification == "AMYLOID POSITIVE":
        print(f'{"║"}  Consider referral for anti-amyloid therapy evaluation       {"║"}')
        print(f'{"║"}  (lecanemab, donanemab). Confirmatory PET optional.          {"║"}')
    elif classification == "AMYLOID NEGATIVE":
        print(f'{"║"}  No evidence of significant amyloid pathology.               {"║"}')
        print(f'{"║"}  Reassure patient. Retest in 1-2 years if risk factors.      {"║"}')
    else:
        print(f'{"║"}  Blood result is indeterminate. Recommend:                   {"║"}')
        print(f'{"║"}  1. Retinal fundus imaging (iFundus VSD) for structural      {"║"}')
        print(f'{"║"}     confirmation of neurodegeneration                        {"║"}')
        print(f'{"║"}  2. If retinal also indeterminate, refer for amyloid PET     {"║"}')

    print(f'{"╚" + "═"*66 + "╝"}')

    return {
        'probability': probability,
        'classification': classification,
        'activations': dict(zip(FEATURE_NAMES, activations.tolist())),
        'raw_values': dict(zip(FEATURE_NAMES, raw_values.tolist())),
    }


# ── Demo: Run report on example patients ────────────────────────────
print('\n── Example 1: Likely AD Patient ──')
r1 = generate_clinical_report(
    pt217_pgml=0.85,    # High pTau217
    ab42_40_ratio=0.05,  # Low Aβ42/40 (amyloid trapped in brain)
    nfl_pgml=25.0,       # Moderate NfL
    gfap_pgml=250.0,     # Elevated GFAP
    patient_name="Example Patient A",
    patient_age=72,
)

print('\n── Example 2: Healthy Control ──')
r2 = generate_clinical_report(
    pt217_pgml=0.08,     # Low pTau217
    ab42_40_ratio=0.10,  # Normal Aβ42/40
    nfl_pgml=12.0,       # Normal NfL
    gfap_pgml=100.0,     # Normal GFAP
    patient_name="Example Patient B",
    patient_age=68,
)

print('\n── Example 3: Non-AD Neurodegeneration (FTD pattern) ──')
r3 = generate_clinical_report(
    pt217_pgml=0.10,     # Normal pTau217 (no amyloid)
    ab42_40_ratio=0.09,  # Normal Aβ42/40
    nfl_pgml=85.0,       # Very high NfL (active neurodegeneration)
    gfap_pgml=180.0,     # Moderate GFAP
    patient_name="Example Patient C",
    patient_age=65,
)


# ══════════════════════════════════════════════════════════════════════
# CELL 8: Manuscript Figures
# ══════════════════════════════════════════════════════════════════════
# %% [markdown]
# ## Step 7: Publication-Quality Figures
# Generate all manuscript figures at 300 DPI with PDF output.

# %%
print('═' * 70)
print('STEP 7: GENERATING MANUSCRIPT FIGURES')
print('═' * 70)

matplotlib.rcParams.update({
    'figure.dpi': 300, 'font.size': 8, 'font.family': 'sans-serif',
    'axes.linewidth': 0.5,
})

# ── Figure 1: Main Results Panel ────────────────────────────────────
fig = plt.figure(figsize=(7.5, 7))
gs = fig.add_gridspec(2, 2, hspace=0.35, wspace=0.35)

# Panel A: ROC
ax = fig.add_subplot(gs[0, 0])
ax.text(-0.18, 1.05, 'A', transform=ax.transAxes, fontsize=14, fontweight='bold')

# Full-data ROCs for display
for name, color, ls, lw in [
    ('pT217', '#9E9E9E', '--', 1.5),
    ('AB42_40', '#EF6C00', ':', 1.0),
    ('VSD_fusion', '#D32F2F', '-', 2.5),
]:
    if name in FEATURE_NAMES:
        ci = FEATURE_NAMES.index(name)
        scores = Ca[:, ci]
        auc = roc_auc_score(y, scores)
        if auc < 0.5: scores = -scores; auc = 1 - auc
    else:
        scores = vsd_scores_full
        auc = auc_full

    fpr_r, tpr_r, _ = roc_curve(y, scores)
    label_map = {'pT217': 'pTau217', 'AB42_40': 'Aβ42/40', 'VSD_fusion': 'VSD 4-channel'}
    ax.plot(fpr_r, tpr_r, color=color, ls=ls, lw=lw,
            label=f'{label_map.get(name, name)} ({auc:.3f})')

ax.plot([0,1],[0,1],'k-',alpha=0.1,lw=0.5)
ax.set_xlabel('1 − Specificity'); ax.set_ylabel('Sensitivity')
ax.legend(fontsize=6.5, loc='lower right')
ax.set_title('Blood biomarkers → amyloid PET', fontsize=9, fontweight='bold')

# Panel B: Method comparison
ax = fig.add_subplot(gs[0, 1])
ax.text(-0.18, 1.05, 'B', transform=ax.transAxes, fontsize=14, fontweight='bold')

plot_methods = method_order
labels_p = [method_labels[m] for m in plot_methods]
means_p = [summary[m]['mean'] for m in plot_methods]
stds_p = [summary[m]['std'] for m in plot_methods]
colors_p = ['#9E9E9E']*4 + ['#EF6C00', '#D32F2F', '#1565C0']

ax.barh(range(len(plot_methods)), means_p, xerr=stds_p, color=colors_p,
        edgecolor='white', lw=0.5, height=0.55, capsize=2, error_kw={'lw':0.7})
ax.set_yticks(range(len(plot_methods)))
ax.set_yticklabels(labels_p, fontsize=7)
ax.set_xlabel('AUC (10×5-fold CV)'); ax.set_xlim(0.55, 0.95)
ax.set_title('Method comparison', fontsize=9, fontweight='bold')
for i, (m, s) in enumerate(zip(means_p, stds_p)):
    ax.text(min(m + s + 0.005, 0.945), i, f'{m:.3f}', fontsize=6.5, va='center')
ax.invert_yaxis()

# Panel C: Score distributions
ax = fig.add_subplot(gs[1, 0])
ax.text(-0.18, 1.05, 'C', transform=ax.transAxes, fontsize=14, fontweight='bold')
bins = np.linspace(0, 1, 50)
ax.hist(vsd_scores_full[y==0], bins=bins, alpha=0.65, color='#1565C0',
        label=f'Amyloid− (n={n_neg})', density=True, edgecolor='white', lw=0.3)
ax.hist(vsd_scores_full[y==1], bins=bins, alpha=0.65, color='#D32F2F',
        label=f'Amyloid+ (n={n_pos})', density=True, edgecolor='white', lw=0.3)
ax.axvline(OPT_THRESHOLD, color='#333', ls='--', lw=1.2)
ax.set_xlabel('VSD fusion score'); ax.set_ylabel('Density')
ax.set_title('Score distributions', fontsize=9, fontweight='bold')
ax.legend(fontsize=7)

# Panel D: Operating point trade-offs
ax = fig.add_subplot(gs[1, 1])
ax.text(-0.18, 1.05, 'D', transform=ax.transAxes, fontsize=14, fontweight='bold')
ax.plot(tpr_f, 1-fpr_f, color='#D32F2F', lw=2)
ax.scatter([OPT_SENS], [OPT_SPEC], color='#D32F2F', s=60, zorder=5,
           edgecolors='black', lw=0.8)
ax.annotate(f'Youden\nSens={OPT_SENS:.0%}\nSpec={OPT_SPEC:.0%}',
            (OPT_SENS, OPT_SPEC), xytext=(OPT_SENS-0.15, OPT_SPEC-0.12),
            fontsize=6.5, arrowprops=dict(arrowstyle='->', color='#666', lw=0.8))
ax.set_xlabel('Sensitivity'); ax.set_ylabel('Specificity')
ax.set_xlim(0.5, 1.02); ax.set_ylim(0.5, 1.02)
ax.set_title('Operating point trade-offs', fontsize=9, fontweight='bold')

plt.tight_layout()
plt.savefig(f'{OUT_DIR}/Figure1_Blood_VSD.png', dpi=300, bbox_inches='tight')
plt.savefig(f'{OUT_DIR}/Figure1_Blood_VSD.pdf', bbox_inches='tight')
plt.show()
print('[SAVED] Figure 1')

# ── Figure 2: Supplementary ─────────────────────────────────────────
fig, axes = plt.subplots(1, 4, figsize=(7.5, 2.2))

# Correlation matrix
ax = axes[0]
ax.text(-0.2, 1.12, 'A', transform=ax.transAxes, fontsize=12, fontweight='bold')
corr = np.corrcoef(Ca.T)
im = ax.imshow(corr, cmap='RdBu_r', vmin=-1, vmax=1, aspect='equal')
ax.set_xticks(range(4)); ax.set_yticks(range(4))
ax.set_xticklabels(FEATURE_NAMES, fontsize=5.5, rotation=45, ha='right')
ax.set_yticklabels(FEATURE_NAMES, fontsize=5.5)
for i in range(4):
    for j in range(4):
        ax.text(j, i, f'{corr[i,j]:.2f}', ha='center', va='center', fontsize=5.5,
                color='white' if abs(corr[i,j]) > 0.55 else 'black')
ax.set_title('Channel correlations', fontsize=7, fontweight='bold')

# Channel weights
ax = axes[1]
ax.text(-0.2, 1.12, 'B', transform=ax.transAxes, fontsize=12, fontweight='bold')
coefs = lr_full.coef_[0]
ch_colors = [CHANNEL_CONFIG[n]['color'] for n in FEATURE_NAMES]
ax.bar(range(4), coefs, color=ch_colors, edgecolor='white', lw=0.5, width=0.6)
ax.set_xticks(range(4))
ax.set_xticklabels(FEATURE_NAMES, fontsize=5.5, rotation=45, ha='right')
ax.set_ylabel('β coefficient', fontsize=6.5)
ax.axhline(0, color='gray', lw=0.5)
ax.set_title('Channel weights', fontsize=7, fontweight='bold')

# Confusion matrix
ax = axes[2]
ax.text(-0.2, 1.12, 'C', transform=ax.transAxes, fontsize=12, fontweight='bold')
cm = confusion_matrix(y, preds_opt)
ax.imshow(cm, cmap='Blues', aspect='equal')
ax.set_xticks([0,1]); ax.set_yticks([0,1])
ax.set_xticklabels(['Pred−','Pred+'], fontsize=6)
ax.set_yticklabels(['True−','True+'], fontsize=6)
for i in range(2):
    for j in range(2):
        ax.text(j, i, f'{cm[i,j]}', ha='center', va='center', fontsize=10, fontweight='bold',
                color='white' if cm[i,j] > cm.max()/2 else 'black')
ax.set_title(f'Confusion matrix', fontsize=7, fontweight='bold')

# Channel scatter
ax = axes[3]
ax.text(-0.2, 1.12, 'D', transform=ax.transAxes, fontsize=12, fontweight='bold')
ax.scatter(Ca[y==0, 0], Ca[y==0, 1], c='#1565C0', alpha=0.2, s=4, rasterized=True)
ax.scatter(Ca[y==1, 0], Ca[y==1, 1], c='#D32F2F', alpha=0.2, s=4, rasterized=True)
ax.set_xlabel('pTau217', fontsize=6); ax.set_ylabel('Aβ42/40', fontsize=6)
ax.set_title('Channel space', fontsize=7, fontweight='bold')

plt.tight_layout()
plt.savefig(f'{OUT_DIR}/Figure2_Supplementary.png', dpi=300, bbox_inches='tight')
plt.savefig(f'{OUT_DIR}/Figure2_Supplementary.pdf', bbox_inches='tight')
plt.show()
print('[SAVED] Figure 2')


# ══════════════════════════════════════════════════════════════════════
# CELL 9: Save All Results
# ══════════════════════════════════════════════════════════════════════
# %%
print('═' * 70)
print('SAVING ALL RESULTS')
print('═' * 70)

output = {
    'title': 'Blood-Based VSD for Alzheimer Amyloid Detection',
    'patent': 'US Provisional 64/038,765',
    'dataset': 'ADNI (Alzheimer Disease Neuroimaging Initiative)',
    'n_participants': int(N),
    'n_amyloid_positive': int(n_pos),
    'n_amyloid_negative': int(n_neg),
    'ground_truth': 'UC Berkeley Amyloid PET (centiloids)',
    'plasma_assays': {
        'Fujirebio_Lumipulse': ['pTau217', 'Aβ42', 'Aβ40'],
        'Quanterix_Simoa': ['NfL', 'GFAP'],
    },
    'vsd_parameters': {
        'sigmoid_steepness': ALPHA,
        'n_channels': 4,
        'channels': {name: {
            'biological_axis': CHANNEL_CONFIG[name]['vsd_channel'],
            'direction': CHANNEL_CONFIG[name]['direction'],
            'individual_AUC': round(ch_aucs[name], 3),
            'calibration_weight': round(float(lr_full.coef_[0][i]), 3),
        } for i, name in enumerate(FEATURE_NAMES)},
    },
    'cross_validation': {
        'method': '10x5-fold repeated stratified',
        'n_folds': 50,
        'results': {method_labels[m]: {
            'AUC_mean': round(summary[m]['mean'], 3),
            'AUC_std': round(summary[m]['std'], 3),
        } for m in method_order},
    },
    'operating_points': {
        'youden_optimal': {
            'threshold': round(float(OPT_THRESHOLD), 4),
            'sensitivity': round(float(OPT_SENS), 3),
            'specificity': round(float(OPT_SPEC), 3),
            'PPV': round(float(PPV), 3),
            'NPV': round(float(NPV), 3),
        },
    },
    'calibration_constants': CALIBRATION,
}

json_path = f'{OUT_DIR}/vsd_blood_complete_results.json'
with open(json_path, 'w') as f:
    json.dump(output, f, indent=2)
print(f'  [SAVED] {json_path}')

# Save calibration table separately (for deployment)
cal_path = f'{OUT_DIR}/vsd_calibration_table.json'
cal_table = {
    'description': 'VSD Blood AD Detection Calibration Table',
    'usage': 'Apply to any new plasma sample with the 4 biomarkers',
    'normalization': CALIBRATION,
    'sigmoid_steepness': ALPHA,
    'channel_weights': {FEATURE_NAMES[i]: round(float(lr_full.coef_[0][i]), 4)
                        for i in range(4)},
    'intercept': round(float(lr_full.intercept_[0]), 4),
    'threshold_youden': round(float(OPT_THRESHOLD), 4),
    'threshold_high_confidence': 1 - CONF_THR,
    'threshold_low_confidence': CONF_THR,
}
with open(cal_path, 'w') as f:
    json.dump(cal_table, f, indent=2)
print(f'  [SAVED] {cal_path}')

print(f'\n{"═"*70}')
print('PIPELINE COMPLETE')
print(f'{"═"*70}')
print(f'''
  ┌─────────────────────────────────────────────────────┐
  │  BLOOD-BASED VSD RESULTS SUMMARY                    │
  ├─────────────────────────────────────────────────────┤
  │  Dataset:     ADNI (n = {N:,})                       │
  │  Ground truth: Amyloid PET ({n_pos} Amy+, {n_neg} Amy−)      │
  │                                                     │
  │  Best method: VSD 4-channel fusion                  │
  │  AUC:         {summary["VSD_fusion"]["mean"]:.3f} ± {summary["VSD_fusion"]["std"]:.3f}                          │
  │  Sensitivity: {OPT_SENS:.1%}                                │
  │  Specificity: {OPT_SPEC:.1%}                                │
  │  NPV:         {NPV:.1%}                                │
  │                                                     │
  │  Key finding: NfL negative weight (β = {lr_full.coef_[0][2]:+.1f})         │
  │  → Disease exclusion signal for non-AD              │
  │                                                     │
  │  Patent: 64/038,765                                 │
  │  Figures: {OUT_DIR}  │
  └─────────────────────────────────────────────────────┘
''')
```
